# The Value of Rapid Antigen Tests to Identify Carriers of Viable SARS-CoV-2

**DOI:** 10.1101/2021.03.10.21252667

**Authors:** Elena V. Shidlovskaya, Nadezhda A. Kuznetsova, Elizaveta V. Divisenko, Maria A. Nikiforova, Andrei E. Siniavin, Daria A. Ogarkova, Aleksandr V. Shagaev, Maria A. Semashko, Artem P. Tkachuk, Olga A. Burgasova, Vladimir A. Gushchin

## Abstract

The search for effective methods to detect patients who excrete a viable virus is one of the urgent tasks of modern biomedicine. In the present study, we examined the diagnostic value of two antigen tests BIOCREDIT COVID-19 Ag (RapiGEN Inc., Korea) and SGTI-flex COVID-19 Ag (Sugentech Inc., Korea) for their diagnostic value in identifying patients who excrete viable SARS-CoV-2. As part of the study, we examined samples from 106 patients who had just been admitted to the hospital, who had undergone quantitative RT-PCR and assessment of viability of SARS-CoV-2 using cell culture. Sensitivity was 0.786 (0.492–0.953) for SGTI-flex COVID-19 Ag and 1 (0.768– 1) for Biocredit COVID-19 Ag. Specificity of rapid tests was significantly higher than that of RT-PCR and was 0.663 (0.557–0.758) and 0.674 (0.568–0.768) for SGTI-flex COVID-19 Ag and Biocredit COVID-19 Ag versus 0.304 (0.213–0.409) obtained for PCR. Thus, for tasks of identifying viable SARS-CoV-2 during screening of conditionally healthy people, as well as monitoring those quarantined, rapid tests show significantly better results.

## Introduction

The SARS-CoV-2 virus pandemic has been a major global problem for over a year. The lack of effective and widely available means of prevention and etiotropic treatment has led to a situation where wearing a mask and social distancing [1] remain the primary ways to lift pressure off the healthcare system, allowing the most severe COVID-19 patients to receive timely and necessary care in medical institutions, while patients with mild to moderate course are forced to remain on lockdown. At the same time, massive restrictions significantly reduce economic activity and, as a result, increase the risks of slowing down economic growth [2].

The main problem of monitoring and surveillance of SARS-CoV-2 is the ability of the virus to spread from asymptomatic patients several days before any symptoms occur [3], [4]. Moreover, contribution to the transmission of the virus from asymptomatic patients and from patients before the onset of symptoms is a significant problem both for the spread of the virus and for accounting for COVID-19 cases [5].

The recent successful launch of SARS-CoV-2 vaccines gives hope for an early reduction of the pandemic and a return to pre-quarantine living conditions [6], [7], [8], [9]. All major vaccines ensure a convincing level of protection (over 90%) in the short term and reliable protection against the severe course of COVID-19. At the same time, the results of the study do not guarantee protection of those vaccinated from subsequent infection with SARS-CoV-2, an asymptomatic course of the disease, which means that further participation of those vaccinated in the spread of the virus has yet to be investigated. The emergence of new strains capable of partial or complete escape from the neutralizing effect of antibodies poses the most danger during prolonged mass vaccination [10], [11], [12].

Detection of viral RNA does not always mean that a person is an infection carrier and spreader. However, in the light of objective epidemic control, it is important to specifically identify carriers of SARS-CoV-2. Timely and prompt identification of the spreaders of infection and their isolation can improve the effectiveness of anti-epidemic measures. Virus viability, as evaluated using cell culture, for samples with a viral load of 30 cycles (about 10^5^–10^6^ GE/mL and below), in RT-PCR tests, is only 3% [13]. It is obvious that the use of methods to assess virus viability using cell culture is unsuitable for mass use due to the complexity of the procedure and the high cost. This requires the search for new simpler methods to identify the spread of the infection.

Rapid antigen tests, which have recently become widespread for the diagnosis of COVID-19, in contrast to PCR, detect SARS-CoV-2 antigens, which, like RNA, comprise viral particles and are produced in infected cells during the life cycle of the virus. The disadvantage of antigen tests is their lower sensitivity compared to RT-PCR. According to the results of Cochrane meta-analysis, sensitivity varies greatly: average sensitivity is 56.2% (95% CI, from 29.5 to 79.8%), average specificity is 99.5% (95% CI, from 98.1% to 99.9%; based on 8 experiments in 5 studies on 943 samples) [14]. At the same time, the definitive advantage of rapid antigen tests is the significantly less laborious process and the ease of learning the procedure, allowing to use the test at home as Point of Care (POC) testing. The time it takes to obtain the result is also crucial, which can be as low as 5 minutes. Moreover, rapid antigen tests are not susceptible to contamination with amplification products, characteristic of nucleic acid analysis methods, which reduces the likelihood of a false positive result.

To date, there are no data on the effectiveness of rapid antigen tests for identification of patients excreting viable SARS-CoV-2. In this paper, we describe results of the pilot study of two rapid antigen tests BIOCREDIT COVID-19 Ag (RapiGEN Inc., Korea) and SGTI-flex COVID-19 Ag (Sugentech Inc., Korea) for their value in identifying patients excreting viable SARS-CoV-2. As part of the study, we examined samples from 106 patients who had just been admitted to the hospital, who had undergone two rapid tests: quantitative RT-PCR and viability assessment of SARS-CoV-2 using susceptible cell culture 293T/ACE2. Samples were collected from January 25, 2021 to February 8, 2021 at an infectious diseases hospital in Moscow.

## Results

### Study design

To investigate the ability of rapid antigen tests to identify patients who excrete viable SARS-CoV-2, it was planned to include primarily patients with suspected COVID-19 newly admitted to the hospital. Participants were admitted to the hospital on 2 to 10 days from the onset of symptoms. The main symptoms were fever, dry cough, chest pain and discomfort, shortness of breath, loss of smell and taste. All included patients had CT signs of lung damage. The study included 106 patients aged 28 to 95 years (mean age 67.67), including 53 women (mean age 68.45) and 53 men (mean age 66.89) (Appendix Table 1).

### Analytical characteristics of antigen tests compared to RT-PCR

Result of PCR tests of smears from 106 patients was positive in 73.58% patients (78 people). The viral load was determined for all positive samples (Appendix Tables 1, 2 and 3), which ranged from 88 to 3.5×10^8^ GE/mL (median 4.6×10^4^). The SGTI-flex COVID-19 Ag rapid test identified 41 of 78 positive samples. Sensitivity of SGTI-flex COVID-19 Ag was 0.526 (95% confidence interval 0.409–0.6399), specificity was 0.964 (0.817–0.999) (Table 1). In turn, the Biocredit COVID-19 Ag rapid test identified 44 of 78 positive samples. For the Biocredit COVID-19 Ag test, sensitivity was 0.564 (0.447– 0.676) and 1.000 (0.877–1.000), respectively. There was no significant differences between the analytical characteristics of the tests (p = 0.8026, McNemar’s test with Edwards continuity correction).

**Table 1.**
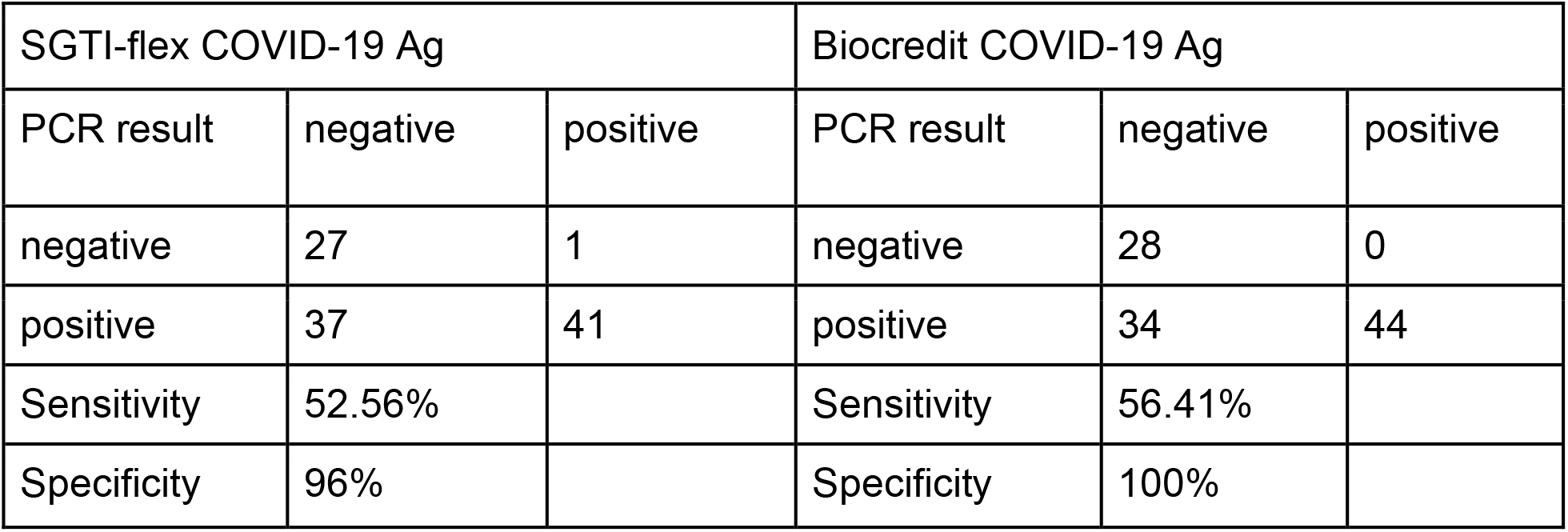
Assessment of sensitivity and specificity of rapid tests compared to RT-PCR.

For samples with a higher viral load, the sensitivity of tests was higher (Appendix Table 2) and starting from a viral load of 1.02E+05 (log10 = 5.0086) for SGTI-flex COVID-19 Ag and 4.74E+04 (log10 = 4.6758) for RapiGen Biocredit COVID-19 Ag, did not differ statistically from the results of PCR tests at a significance level of 0.01 (p = 0.01333, McNemar’s test with Edwards continuity correction). P-value only increased with further increase in viral load.

To determine the analytical threshold of sensitivity with respect to the antigen in virions of the culture fluid, we conducted a model experiment, when the culture fluid with a known virus titer was used to assess the analytical sensitivity of rapid tests in the range from 10^2^ to 10^8^ GE/mL, using an interval of one order of magnitude. Both tests showed the detection limit at a virus titer of 10^6^ GE/mL (10^5^ GE/test), which corresponded to 4×10^5^ TCID50/mL or 4×10^4^ TCID50/test. There was no statistically significant difference in analytical characteristics depending on the day from the onset of the disease (p-value = 0.2356 for SGTI-flex COVID-19 Ag, p-value = 0.8581 for RapiGen Biocredit COVID-19 Ag with Fisher’s exact test), which is not surprising given that there were high-load patients on each day (Appendix Table 3).

### Analytical characteristics of antigen tests for viability

All samples were evaluated for viability of SARS-CoV-2 virus using a sensitive cell culture. Viability was assessed using the 293T/ACE2 cell line with stable expression of the human ACE2 receptor. For samples with a cytopathogenic effect (CPE), RT-PCR was performed to confirm that the CPE was caused by SARS-CoV-2 and not by other infectious agents. Comparison of groups of samples with viable and non-viable viral load measured by quantitative PCR showed a significant difference (p<0.0001, p-value calculated using the Mann–Whitney test) (Fig. 1).

**Figure 1.**
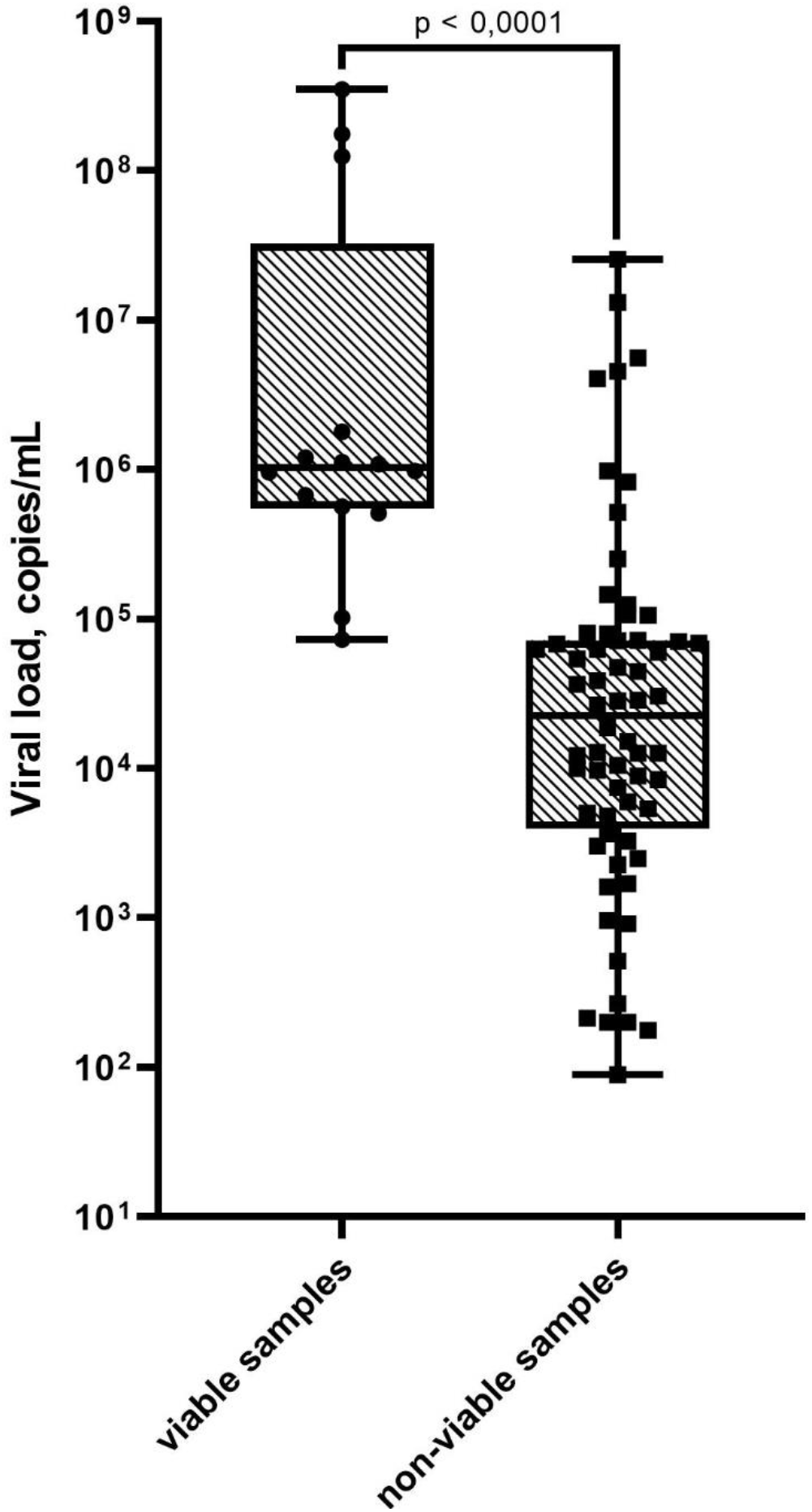
SARS-CoV-2 viral load in viable versus non-viable samples. Results are represented by a box plot: horizontal lines – medians; boxes – interquartile range; whiskers – min-max (p-value calculated using Mann–Whitney test)

Viability was shown only by samples with a viral load of 7.3×10^4^ (GE/mL) and higher (Fig. 1 and Table 2). However, not all samples with such a load remained viable. Rapid tests were able to give a positive result on samples with median values of 5.72×10^4^ and 3.78×10^4^ (GE/mL) for Biocredit COVID-19 Ag and SGTI-flex COVID-19 Ag, respectively. However, a positive result was also obtained for a number of samples with a load below 10^4^ (GE/mL). It is not clear whether this is related to how the material was collected for three different tests or the presence in some biological samples of a disproportionate amount of antigen in relation to viral RNA.

**Table 2.** Viral load (GE/mL) depending on viability and rapid test result.

| Viral load (GE/mL) |  |  |  |  |
| --- | --- | --- | --- | --- |
| quantitative real-time RT-PCR |  |  |  |  |
| value | Number of positive tests | Viral load (mean) | range | median |
| Successful isolation | 14 | $4.71 \times 10^7$ | $7.30 \times 10^4 - 3.50 \times 10^8$ | $1.03 \times 10^6$ |
| Unsuccessful isolation | 64 | $8.93 \times 10^5$ | $89.00 - 2.56 \times 10^7$ | $2.26 \times 10^4$ |
| BIOCREDIT COVID-19 Ag |  |  |  |  |
| Successful isolation | 14 | $4.71 \times 10^7$ | $7.30 \times 10^4 - 3.50 \times 10^8$ | $1.03 \times 10^6$ |
| Unsuccessful isolation | 30 | $1.87 \times 10^6$ | $89.00 - 2.56 \times 10^7$ | $5.72 \times 10^4$ |
| RT-PCR “+”,<br>antigen test “-” | 34 | $2.87 \times 10^4$ | $1.77 \times 10^2 - 1.46 \times 10^5$ | $9.85 \times 10^3$ |
| SGTI-flex COVID-19 Ag |  |  |  |  |
| Successful isolation | 11 | $5.97 \times 10^7$ | $7.30 \times 10^4 - 3.50 \times 10^8$ | $1.09 \times 10^6$ |
| Unsuccessful isolation | 30 | $1.40 \times 10^6$ | $89.00 - 2.56 \times 10^7$ | $3.78 \times 10^4$ |
| RT-PCR “+”,<br>antigen test “-” | 37 | $4.87 \times 10^5$ | $1.77 \times 10^2 - 1.31 \times 10^7$ | $1.87 \times 10^4$ |

Overall, out of 106 samples, viable virus was detected in 14 patients, representing 13.2% of all participants. Using these samples, SGTI-flex COVID-19 Ag gave 11 (78.6%) positive results, while Biocredit COVID-19 Ag gave 14 (100%) positive results (Table 3). As a result, the sensitivity of antigen tests was 0.786 (0.492–0.953) for SGTI-flex COVID-19 Ag and 1 (0.768–1) for Biocredit COVID-19 Ag. Specificity was 0.663 (0.557–0.758) and 0.674 (0.568–0.768) for SGTI-flex COVID-19 Ag and Biocredit COVID-19 Ag, respectively. For RT-PCR, the sensitivity was 1 (0.768–1), while the specificity was 0.304 (0.213–0.409).

**Table 3.**
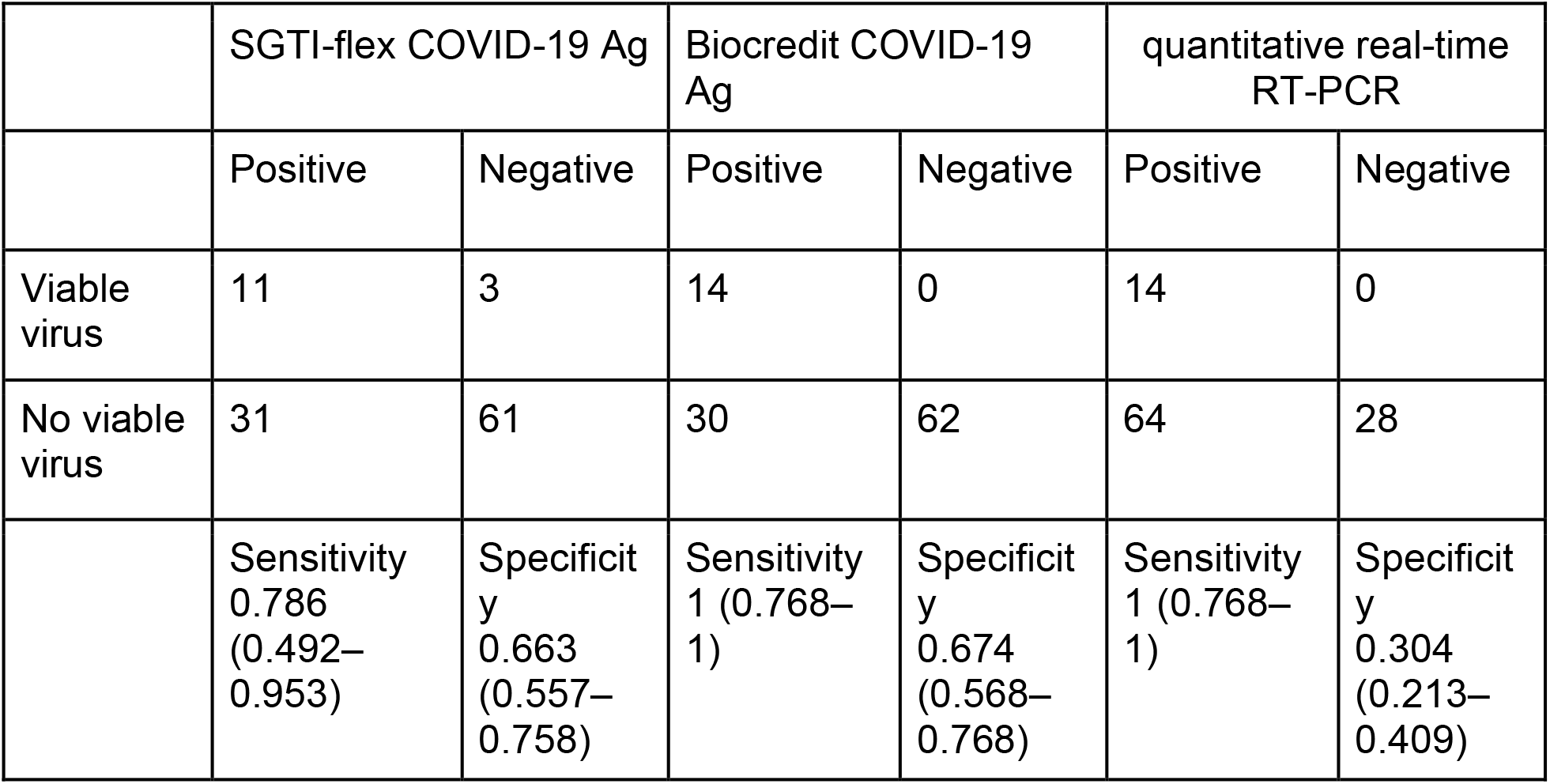
Assessment of sensitivity and specificity of rapid tests for viability.

Comparing analytical characteristics of the tests for viability of the virus, it can be observed that the rapid tests are indistinguishable from each other (p-value = 0.4533, McNemar’s test with Edwards continuity correction), but they differ significantly from RT-PCR (p-value = 2.546e-06 for SGTI-flex COVID-19 Ag, p-value = 1.519e-08 for Biocredit COVID-19 Ag, McNemar test with Edwards continuity correction).

As the viral load increased, the sensitivity of the tests for viral viability increased and, starting from a viral load of 6.27E+04 (log10 = 4.7973) for SGTI-flex COVID-19 Ag and 5.11E+05 (log10 = 5.7084) for RapiGen Biocredit COVID-19 Ag, did not statistically significantly differ from the possibility of successful isolation of a viable virus at a significance level of 0.01 (p = 0.01529 and p = 0.01333, respectively).

There were no statistical differences in the analytical characteristics of rapid tests depending on the day of the disease (p-value = 0.5292 for SGTI-flex COVID-19 Ag, 0.4108 for RapiGen Biocredit COVID-19 Ag using Fisher’s exact test). At the same time, the number of cases of successful isolation of a viable virus also did not statistically differ at different time intervals of the disease, which is probably due to the limited follow-up period and the use of only one sample from each patient.

## Discussion

Prompt identification of carriers of the viable SARS-CoV-2 virus and their timely isolation from the society is the most important step in containing a pandemic. Mass screening, in turn, is limited by access of the population to laboratory testing, which in the case of PCR is limited by the capabilities of laboratories. In this regard, many states are adopting new approaches, whereby samples are pooled to increase the amount of PCR tests [15], [16], [17], [18].

An alternative to using PCR laboratories is to use rapid antigen tests, including for home use. In December 2020, the FDA for the first time authorized home use of an antigen test to detect the virus [19]. There is no doubt that moving in this direction can significantly improve access to testing. Evaluation of cost-effectiveness of mass screening shows that in the case of, for example, the USA, the most optimal strategy is weekly testing and two-week isolation of the infected person or monthly testing with weekly isolation for scenarios of rapid (Re = 2.2) and slow (Re = 1.2) epidemic, respectively [20]. The important point is that the use of antigen tests can significantly increase the effectiveness of regular mass testing.

Recommendations regarding self-isolation and lockdown deserve special attention. The existing WHO recommendations prescribe to revoke termination of isolation for patients with manifest forms 13 days after the onset of symptoms, provided that in the last 3 days the person did not experience any characteristic symptoms of the disease, as well as 10 days after a positive PCR result for asymptomatic cases [21]. Those who have come into contact with a person with a confirmed diagnosis of COVID-19 are also prescribed a 14-day quarantine, however testing during the quarantine or upon its termination is not mandatory. Currently such a strategy is subject to serious scientific criticism [22], [23]. Re-testing at the beginning of quarantine and upon its termination, but with a shorter time spent in isolation, as well as daily testing instead of quarantine, according to the results of a mathematical model, can be more effective in protecting against the spread of the infection after the quarantine is over. An additional benefit of a shorter quarantine is that the person returns to work faster, which reduces the economic damage of the pandemic lock down. It is worth noting that for the purposes of mathematical modeling, a patient with a positive test result is considered potentially infectious to others. In this connection, using tests that improve the accuracy of the evaluation of the number of persons excreting the virus will allow to further improve the effectiveness of monitoring.

WHO notes that studies to evaluate the viability of the virus in patient samples are currently very limited. Viable virus was excreting in persons without symptoms, in patients with mild to moderate course of COVID-19 for 8–9 days after the onset of symptoms, and for a longer time period in critical patients [21]. However, a meta-analysis of the results of isolation of RNA and the viable virus extraction shows that although SARS-CoV-2 RNA is excreted for several weeks, viable virus is not detected already after 9 days [24]. Thus, given the significant difference in the dynamics of isolation of RNA and the viable virus, there is now the need to look for tests that allow to identify persons who excrete viable virus while minimizing the likelihood of a false-positive result in patients who no longer excrete viable virus. This will minimize the amount of persons quarantined, reducing the negative economic damage of the pandemic. It also allows epidemiological services to focus on patients who are the real sources of infection spread. In this study, we investigated the value of antigen tests for identifying carriers of a viable virus.

We examined samples from 106 patients admitted to a hospital in Moscow. The study included only those patients who had experienced first symptoms no earlier than 10 days before. All samples, in addition to being used for the two tests, were also used for quantitative PCR and assessment of viral viability using cell culture. Of 106 samples, 78 (73.58%) were PCR-positive. This is significantly higher than previously published data with similar clinical setting [25]. This is probably due to a difference in design, as for the purposes of this study, we searched for patients who had just been admitted to the hospital, while the other studies did not make such a distinction [26], [27], [28].

Data of quantitative PCR are of particular interest. The viral load was determined for all PCR-positive samples and it was found that it varied greatly, from 88 to 3.5×10^8^ GE/mL (median 4.6×10^4^). In terms of viral load, samples that showed viability were significantly different from the rest (p<0.0001, the p-value was calculated using the Mann–Whitney test). For all 14 samples that showed viability, the viral load was at least 7.3×10^4^ (GE/mL). And although not all samples with a similar load or higher showed viability, it is important that a cut-off quantitative threshold, after which the probability of the virus remaining viable is greatly reduced, can be established experimentally.

Standard evaluation of analytical characteristics of rapid tests showed that, relative to RT-PCR, their diagnostic characteristics were close to the mean values published for the Cochrane meta-analysis [14]. For the SGTI-flex COVID-19 Ag test, sensitivity was 0.526 (95% confidence interval 0.409–0.6399), specificity was 0.964 (0.817–0.999). For the Biocredit COVID-19 Ag test, sensitivity and specificity were 0.564 (0.447–0.676) and 1.000 (0.877–1.000), respectively. No significant differences were found between tests (p = 0.8026). Analysis of the value of tests to detect samples containing viable virus showed that both tests are highly sensitive. Sensitivity was 0.786 (0.512–0.942) and 1 (0.755-1) for SGTI-flex COVID-19 Ag and Biocredit COVID-19 Ag. The control method (RT-PCR) had sensitivity of 1 (0.886–1). In turn, specificity of rapid tests was significantly higher than that of RT-PCR and was 0.663 (0.557–0.758) and 0.674 (0.568–0.768) for SGTI-flex COVID-19 Ag and Biocredit COVID-19 Ag versus 0.304 (0.213–0.409) obtained for PCR. Statistically, the results of the antigen tests used in the study are indistinguishable from each other (p-value = 0.4533), but differ significantly from RT-PCR (p-value = 2.546e-06 for SGTI-flex COVID-19 Ag, p-value = 1.519e-08 for Biocredit COVID-19 Ag). This means that rapid tests have significantly better results for the task of identifying viable SARS-CoV-2.

We are not aware of any other studies that analyze the value of antigen tests to identify patients who excrete viable virus. Results obtained in this study require confirmation using more samples from patients who excrete viable virus. In this study, there were 14 samples that showed viability. It appears important to investigate to what extent the results of this study will correlate with data obtained on samples from asymptomatic patients. It is known that such patients contribute significantly to the transmission of the virus, which in the context of our study means a higher viral load and, consequently, virus viability of these patients. Further investigation will clarify this issue and help to understand the value of antigen tests for the widespread detection of infectious agents in the context of the COVDI-19 pandemic.

## Conclusion

This article presents the results of a study on the value of rapid antigen tests to identify patients who excrete viable SARS-CoV-2. With a comparable effectiveness of detection of patients excreting a viable virus, antigen tests, relative to RT-PCR, showed significantly fewer false-positive results on patient samples that no longer excrete a viable virus, and therefore are safe for others. This suggests that practical use of antigen tests can lead to shorter isolation time and quarantine of people who are safe for general public.

Undoubtedly, a direct comparison of the analytical characteristics of RT-PCR with antigen tests is not entirely correct, since the sensitivity of RT-PCR significantly exceeds the sensitivity of antigen tests, however, to identify patients excreting a viable virus, the value of antigen tests, according to the result we obtained, is higher. It is important to note that antigen testing does not require sophisticated equipment, trained personnel and complex quality control systems, which means that the widespread use of antigen tests for mass screening and detection of persons excreting the virus among conditionally healthy people is potentially of great value.

## Materials and methods

### Patients

The study included patients with suspected COVID-19 admitted to the hospital on day 2–10 from the onset of symptoms (fever, dry cough, chest pain and discomfort, shortness of breath, loss of smell and taste) and with CT signs of lung damage. Study was approved by the Local ethic committee of the Moscow First Infectious Diseases Hospital (the Protocol #2 dated 2021-01-22). All participants signed the written informed consent to allow usage of nasal swab samples for research purposes.

The study included a total of 106 patients aged 28 to 95 years (mean age 67.67) including 53 women (mean age 68.45) and 53 men (mean age 66.89).

### Sample collection and transportation

Using sterile swabs and observing the necessary safety precautions, nurses of the hospital collected three nasopharyngeal samples from each patient, two of which were used for antigen testing. The third sample was transferred to tubes with 1 mL of phosphate buffered saline (PBS). All collected materials were transferred to the reference center for coronavirus infection of the Gamaleya Research Institute of Epidemiology and Microbiology of the Ministry of Health of Russia cooled down to +4 degrees Celsius within 2 hours after collection.

### Antigen testing

Antigen testing was done immediately after sample collection in accordance with the manufacturer’s instructions directly at the patient’s bedside. Testing was done using two commercially available rapid tests for the detection of SARS-CoV-2 antigen – BIOCREDIT COVID-19 Ag (RapiGEN Inc., Korea) and SGTI-flex COVID-19 Ag (Sugentech Inc., Korea).

### SARS-CoV-2 testing

All collected samples were tested immediately after transportation. PCR amplification was carried out using a one-step “SARS-CoV-2 FRT” commercial kit with catalog number EA-128 (bought from N.F. Gamaleya NRCEM, Moscow, Russia). According to manufacturer’s information “SARS-CoV-2 FRT” kit allows to amplify a fragment from the 5’ end region encoding the NSP1 gene (aprox. 450 to 650 nt bases upstream the 5’ end of SARS-CoV-2 viral genome). The protocol for qPCR-RT used in this study had been described previously [29]. Briefly the conditions of the one-step RT-qPCR reaction were as follows: 50°C for 15 min, 95°C for 5 min, followed by 45 cycles of 95°C for 10 s and 55°C for 1 min. The number of copies of viral RNA was calculated using a standard curve generated by amplification of plasmid cloned DNA template fragment encoding 450 to 650 nt bases upstream the 5’ end of SARS-CoV-2 viral genome.

### Virus isolation

Isolation of the SARS-CoV-2 virus was performed using 293T/ACE2 cell line (with stable expression of the human ACE2 receptor). Cells were cultured in DMEM medium (PanEco, Russia) containing 10% FBS (HyClone, USA), 1% L-glutamine, and 1% penicillin/streptomycin. 96-well plate was used for the experiment. For this, nasopharyngeal secretion (100 μL) from COVID-19 patients was added to tablets. Plates were incubated for 5 days. Virus-induced cytopathic effect (CPE) was then assessed. Also, for samples with CPE, real-time PCR was performed to confirm that CPE was caused by SARS-CoV-2 and not by other infectious agents that can cause CPE.

### Statistical treatment of results

All data were statistically treated using the methods available in different R packages. McNemar’s test with Edwards continual correction was used to compare the two different tests. Fisher’s exact test was used to analyze unrelated qualitative data. Quantitative indicators were checked for normal distribution using the Shapiro–Wilk test. Quantitative comparison of groups was done using the Mann–Whitney test. Confidence intervals for specificity and sensitivity, as well as confidence intervals for proportions, were calculated using binomial distribution with the help of Clopper–Pearson method.

## Data Availability

All data generated or analysed during this study are included in this published article (and its supplementary information files).

