## Supplemental Tables 1, 2, 3 for "The Value of Rapid Antigen Tests to Identify Carriers of Viable SARS-CoV-2"

### Supplementary

**Table 1.** Baseline data for all patients

| ID | Gender | Age range | PCR results (Ct) | Viral load, copies/ ml | SGTI-flex COVID-19 Ag | Biocredit COVID-19 Ag | Days from onset of symptoms | Virus isolation |
|---|---|---|---|---|---|---|---|---|
| 1 | female | 75-79 | 33,83 | 3,65E+04 | neg | neg | 5 | - |
| 2 | male | 75-79 | neg | | neg | neg | 4 | - |
| 3 | female | 45-49 | 28,23 | 5,11E+05 | pos | pos | 3 | + |
| 4 | female | 45-49 | 34,65 | 5,36E+03 | pos | pos | 6 | - |
| 5 | male | 45-49 | 27,46 | 8,26E+05 | neg | pos | 6 | - |
| 6 | female | 70-74 | neg | | neg | neg | 6 | - |
| 7 | female | 30-34 | 35,13 | 1,26E+04 | neg | neg | 6 | - |
| 8 | male | 65-69 | 23,09 | 2,56E+07 | pos | pos | 3 | - |
| 9 | male | 75-79 | neg | | neg | neg | 4 | - |
| 10 | female | 65-69 | neg | | neg | neg | 5 | - |
| 11 | male | 70-74 | neg | | neg | neg | 4 | - |
| 12 | male | 45-49 | 31,92 | 7,07E+04 | pos | pos | 4 | - |
| 13 | male | 50-54 | 33,72 | 2,49E+03 | neg | neg | 5 | - |
| 14 | male | 85-89 | 31,51 | 1,05E+04 | pos | pos | 6 | - |
| 15 | male | 70-74 | neg | | neg | neg | 5 | - |
| 16 | male | 80-84 | 23,25 | 4,05E+06 | pos | pos | 2 | - |
| 17 | female | 65-69 | 23,51 | 1,12E+06 | pos | pos | 2 | + |
| 18 | male | 60-64 | 31,27 | 1,22E+04 | pos | neg | 6 | - |
| 19 | female | 80-84 | 29,18 | 4,74E+04 | pos | pos | 4 | - |
| 20 | female | 80-84 | 28,62 | 6,81E+04 | pos | pos | 5 | - |
| 21 | male | 30-34 | 32,25 | 2,87E+04 | neg | neg | 4 | - |
| 22 | male | 80-84 | neg | | neg | neg | 1 | - |
| 23 | male | 50-54 | neg | | neg | neg | 5 | - |
| 24 | female | 40-44 | neg | | neg | neg | 4 | - |
| 25 | female | 60-64 | neg | | neg | neg | 1 | - |
| 26 | female | 80-84 | 33,72 | 3,27E+03 | neg | neg | 6 | - |
| 27 | female | 70-74 | 29,93 | 7,30E+04 | pos | pos | 6 | + |
| 28 | female | 75-79 | 25,04 | 1,20E+06 | neg | pos | 9 | + |
| 29 | male | 75-79 | 35,51 | 9,62E+02 | pos | neg | 8 | - |
| 30 | female | 75-79 | 25,18 | 1,09E+06 | pos | pos | 6 | + |
| 31 | male | 50-54 | 30,86 | 6,27E+04 | neg | neg | 8 | - |
| 32 | female | 60-64 | 31,52 | 4,46E+04 | neg | neg | 8 | - |
| 33 | female | 90-94 | 29,02 | 7,98E+04 | neg | neg | 8 | - |
| 34 | male | 60-64 | 24,45 | 1,79E+06 | pos | pos | 9 | + |
| 35 | female | 80-84 | 25,89 | 6,72E+05 | pos | pos | 9 | + |
| 36 | male | 60-64 | 26,74 | 9,77E+05 | neg | pos | 10 | + |
| 37 | male | 65-69 | 34,68 | 1,69E+03 | neg | neg | 9 | - |
| 38 | female | 65-69 | 28,59 | 1,07E+05 | neg | neg | 8 | - |
| 39 | female | 80-84 | 30,81 | 2,65E+04 | pos | pos | 3 | - |
| 40 | male | 70-74 | 32,08 | 8,90E+03 | neg | pos | 5 | - |

| | | | | | | | | |
|---|---|---|---|---|---|---|---|---|
| 41 | female | 60-64 | 30,65 | 3,04E+04 | neg | pos | 5 | - |
| 42 | female | 80-84 | 31,46 | 1,51E+04 | neg | pos | 4 | - |
| 43 | male | 80-84 | 34,06 | 1,61E+03 | neg | neg | 5 | - |
| 44 | female | 77-79 | 29,7 | 6,88E+04 | pos | pos | 8 | - |
| 45 | female | 45-49 | 31,21 | 1,87E+04 | neg | neg | 6 | - |
| 46 | male | 45-49 | neg | | neg | neg | 8 | - |
| 47 | male | 80-84 | 20,99 | 1,25E+08 | pos | pos | 2 | + |
| 48 | female | 70-74 | 29,52 | 8,05E+04 | neg | neg | 8 | - |
| 49 | female | 60-64 | 33,66 | 2,27E+03 | neg | neg | 8 | - |
| 50 | female | 75-79 | 19,79 | 3,50E+08 | pos | pos | 10 | + |
| 51 | male | 60-64 | neg | | neg | neg | 7 | - |
| 52 | female | 60-64 | 31,24 | 7,16E+04 | pos | pos | 9 | - |
| 53 | female | 70-74 | neg | | neg | neg | 9 | - |
| 54 | female | 70-74 | neg | | neg | neg | 6 | - |
| 55 | female | 55-59 | neg | | neg | neg | 9 | - |
| 56 | male | 80-84 | 35,8 | 9,97E+03 | pos | neg | 4 | - |
| 57 | female | 65-69 | neg | | neg | neg | 8 | - |
| 58 | male | 80-84 | 24,52 | 1,31E+07 | neg | pos | 8 | - |
| 59 | female | 65-69 | 30,33 | 1,06E+05 | neg | neg | 8 | - |
| 60 | male | 80-84 | 30,43 | 1,02E+05 | pos | pos | 7 | + |
| 61 | male | 70-74 | 32,66 | 3,87E+04 | neg | neg | 9 | - |
| 62 | female | 60-64 | neg | | pos | neg | 8 | - |
| 63 | male | 80-84 | 31,54 | 6,29E+04 | neg | neg | 9 | - |
| 64 | male | 70-74 | 31,89 | 5,41E+04 | neg | pos | 8 | - |
| 65 | female | 75-79 | 29,6 | 1,46E+05 | neg | neg | 9 | - |
| 66 | female | 50-54 | 26,47 | 5,66E+05 | pos | pos | 9 | + |
| 67 | male | 80-84 | neg | | neg | neg | 8 | - |
| 68 | male | 55-59 | 37,48 | 4,80E+03 | neg | neg | 9 | - |
| 69 | male | 55-59 | neg | | neg | neg | 6 | - |
| 70 | male | 60-64 | 33,39 | 2,83E+04 | pos | pos | 8 | - |
| 71 | male | 75-79 | 26,98 | 4,54E+06 | pos | pos | 4 | - |
| 72 | female | 70-74 | 33,43 | 3,66E+03 | neg | neg | 9 | - |
| 73 | female | 70-74 | 33,01 | 5,01E+03 | neg | neg | 6 | - |
| 74 | male | 60-64 | neg | | neg | neg | 7 | - |
| 75 | male | 85-89 | 36,04 | 5,13E+02 | pos | pos | 3 | - |
| 76 | female | 70-74 | 26,02 | 9,59E+05 | neg | pos | 5 | + |
| 77 | female | 85-89 | 35,28 | 9,12E+02 | pos | neg | 3 | - |
| 78 | female | 85-89 | 28,73 | 1,25E+05 | pos | pos | 7 | - |
| 79 | female | 40-44 | 19,09 | 1,76E+08 | pos | pos | 3 | + |
| 80 | male | 55-59 | 37,46 | 1,77E+02 | neg | neg | 4 | - |
| 81 | male | 70-74 | 25,99 | 9,81E+05 | pos | pos | 9 | - |
| 82 | male | 85-89 | 26,83 | 5,19E+05 | pos | pos | 7 | - |
| 83 | male | 50-54 | 35,99 | 2,13E+02 | neg | neg | 4 | - |
| 84 | female | 80-84 | neg | | neg | neg | 9 | - |
| 85 | male | 40-44 | 32,72 | 3,02E+03 | neg | neg | 7 | - |
| 86 | female | 60-64 | neg | | neg | neg | 9 | - |
| 87 | male | 60-64 | neg | | neg | neg | 9 | - |
| 88 | female | 80-84 | 35,71 | 2,68E+02 | neg | neg | 8 | - |

| | | | | | | | | |
|---|---|---|---|---|---|---|---|---|
| 89 | male | 80-84 | neg | | neg | neg | 9 | - |
| 90 | female | 55-59 | 37,07 | 8,87E+01 | pos | pos | 9 | - |
| 91 | male | 50-54 | neg | | neg | neg | 6 | - |
| 92 | female | 60-64 | 36,06 | 2,00E+02 | pos | pos | 9 | - |
| 93 | female | 60-64 | neg | | neg | neg | 7 | - |
| 94 | female | 85-89 | 31,45 | 8,42E+03 | pos | pos | 5 | - |
| 95 | female | 70-74 | 36,06 | 2,01E+02 | pos | neg | 9 | - |
| 96 | female | 80-84 | 29,69 | 6,03E+04 | pos | pos | 8 | - |
| 97 | female | 25-29 | 31,61 | 7,44E+03 | pos | pos | 6 | - |
| 98 | male | 75-79 | 27,26 | 2,52E+05 | pos | pos | 6 | - |
| 99 | male | 60-64 | 31,88 | 5,97E+03 | neg | neg | 3 | - |
| 100 | male | 70-74 | 34,32 | 1,26E+04 | neg | neg | 10 | - |
| 101 | male | 45-49 | 35,56 | 9,74E+03 | neg | neg | 10 | - |
| 102 | female | 80-84 | 32,11 | 7,20E+04 | pos | neg | 6 | - |
| 103 | male | 50-54 | 23,67 | 5,59E+06 | pos | pos | 8 | - |
| 104 | male | 80-84 | neg | | neg | neg | 6 | - |
| 105 | male | 50-54 | neg | | neg | neg | 10 | - |
| 106 | male | 55-59 | 34,3 | 1,28E+04 | pos | pos | 5 | - |

**Table 2.** Distribution of test results by viral load as measured by quantitative RT-PCR

| Viral load | 1.25E+08 – 3.50E+08 | 1.31E+07 – 2.56E+07 | 1.09E+06 – 5.59E+06 | 1.02E+05 – 9.81E+05 | 1.05E+04 – 8.05E+04 | 1.61E+03 – 9.97E+03 | 1.77E+02 – 9.62E+02 | 8.87E+01 |
|---|---|---|---|---|---|---|---|---|
| Number of samples (PCR) | 3 | 2 | 7 | 14 | 27 | 16 | 8 | 1 |
| Sugentech SGTI-flex COVID-19 Ag | 3 (100%) | 1(50%) | 6 (85.71%) | 8 (57.14%) | 13 (48.15%) | 4 (25%) | 5 (62.5%) | 1 (100%) |
| RapiGen Biocredit COVID-19 Ag | 3 (100%) | 2 (100%) | 7 (100%) | 11 (78.57%) | 14 (51.85%) | 4 (25%) | 2 (25%) | 1 (100%) |

**Table 3.** Distribution of test results over time from day 2 to day 10 after the onset of symptoms

| Time since onset of symptoms | 2 days | 3 days | 4 days | 5 days | 6 days | 7 days | 8 days | 9 days | 10 days |
|---|---|---|---|---|---|---|---|---|---|
| Number of positive samples (PCR) | 3 | 7 | 8 | 9 | 12 | 4 | 16 | 15 | 4 |
| Viral load | 1.12E+06<br>1.25E+08<br>4.05E+06 | 1.76E+08<br>2.56E+07<br>2.65E+04<br>5.11E+05<br>5.13E+02<br>5.97E+03<br>9.12E+02 | 1.51E+04<br>1.77E+02<br>2.13E+02<br>2.87E+04<br>4.54E+06<br>4.74E+04<br>7.07E+04<br>9.97E+03 | 1.28E+04<br>1.61E+03<br>2.49E+03<br>3.04E+04<br>3.65E+04<br>6.81E+04<br>8.42E+03<br>8.90E+03<br>9.59E+05 | 1.05E+04<br>1.09E+06<br>1.22E+04<br>1.26E+04<br>1.87E+04<br>2.52E+05<br>3.27E+03<br>5.01E+03<br>5.36E+03<br>7.20E+04<br>7.44E+03<br>8.26E+05 | 1.02E+05<br>1.25E+05<br>3.02E+03<br>5.19E+05 | 1.06E+05<br>1.07E+05<br>1.31E+07<br>2.27E+03<br>2.68E+02<br>2.83E+04<br>4.46E+04<br>5.41E+04<br>5.59E+06<br>6.03E+04<br>6.27E+04<br>6.88E+04<br>7.30E+04<br>7.98E+04<br>8.05E+04<br>9.62E+02 | 1.20E+06<br>1.46E+05<br>1.69E+03<br>1.79E+06<br>2.00E+02<br>2.01E+02<br>3.66E+03<br>3.87E+04<br>4.80E+03<br>5.66E+05<br>6.29E+04<br>6.72E+05<br>7.16E+04<br>8.87E+01<br>9.81E+05 | 1.26E+04<br>3.50E+08<br>9.74E+03<br>9.77E+05 |
| Sugentech SGTI-flex COVID-19 Ag | 3 | 6 | 4 | 3 | 8 | 3 | 5 | 8 | 1 |
| RapiGen Biocredit COVID-19 Ag | 3 | 5 | 4 | 6 | 7 | 3 | 6 | 8 | 2 |
